# Temporal Characterization of Longitudinal Sequelae Including Acute Pain, Physiologic Status, and Toxicity Kinetics in Head and Neck Cancer Patients Receiving Radiotherapy: A Prospective Electronic Health Record Embedded Registry Study

**DOI:** 10.1101/2022.05.05.22274717

**Authors:** Vivian Salama, Sara Youssef, Tianlin Xu, Jaime Chen, Jillian Rigert, Kareem Wahid, Cielito C. Reyes-Gibby, Anna Lee, Katherine A. Hutcheson, Brandon Gunn, David I. Rosenthal, Abdallah S. R. Mohammed, Lisanne V van Dijk, Ruitao Lin, Clifton D. Fuller, Amy C. Moreno

## Abstract

**Background:** Pain is a common, debilitating symptom experienced by patients with oral cavity and oropharyngeal cancer (OC/OPC) treated with radiotherapy (RT). Managing acute pain (AP) over 6 - 7 weeks of RT remains a significant challenge, warranting further investigation. Using a modern prospective registry, the objective of this study was to characterize longitudinal AP profiles and temporal changes in vital signs (VS), radiation toxicities, and analgesic prescribing patterns during RT.

**Methods:** A total of 351 patients with OC (n=120) and OPC (n=228) treated with curative RT from 2013-2021 were included. Baseline cohort characteristics, weekly patient-reported pain descriptors, physician-graded toxicities (CTCAE v5), and analgesic orders during RT were extracted. Temporal changes in AP scores and VS were analyzed using linear mixed effect models. AP trajectories were reduced to single metric area under the curve calculations (AUC*pain*). Correlations were assessed using Spearman correlation coefficients.

**Results:** Median age was 60 years, and 70% and 42% received chemotherapy and surgery, respectively. A significant increase in pain, mucositis, dermatitis, and overall treatment toxicity severity were observed by the end of RT. AUC*pain* was significantly different based on gender, primary tumor site, surgery, drug use history and pre-RT pain. There was a temporal mean weight loss of 7.1% bodyweight (95%CI, 10-8.2; *P*<0.001), a mean arterial pressure (MAP) decline of 6.8 mmHg (95%CI, −8.8 to −4.7; *P*<0.001), and increased pulse rate of 11 beats/min (95%CI, 7.6-13.8; *P*<0.001). AP and pulse rate were positively associated over time (*P<0*.*001*) while weight and MAP were negatively associated over time (*P*<0.001). A temporal increase in analgesics use, mainly opioids, was detected.

**Conclusion:** This study characterizes longitudinal treatment-related toxicity kinetics using a prospective OC/OPC registry and demonstrates an ongoing need for optimized, timely pain control. Pain AUC metrics preserve temporal information and may be useful for developing algorithmic pain prediction and management models.

## Background

Oral cavity and oropharyngeal cancers (OC/OPC) afflict 450,000 patients annually with over 54,000 cases diagnosed in the United States alone.(1)Treatment of OC/OPC is stage and site dependent with locally advanced stages generally treated using a multimodal approach (i.e., surgery, chemotherapy, and/or radiotherapy [RT]) whereas surgery followed by postoperative RT is the standard of care for OC cases with high-risk features. (2) Although advanced novel techniques in various treatment modalities have been shown to improve survival, several acute toxicities experienced during RT can translate into chronic conditions radiation-attributable sequelae, thereby negatively impacting the quality of life of survivors.(3)

Pain is a commonly experienced toxicity that can be present before (i.e., cancer-related) or during and after RT (i.e., treatment-related).(4) The presence of pre-treatment pain, identified as an independent predictor of survival OC/OPC, can make it increasingly challenging to manage pain during therapy.(5) Acute pain (during RT) in the mouth/throat region has been reported to affect over 90% of head and neck cancer (HNC) patients, with 50-80% requiring an opioid prescription. (6–8) While the World Health Organization (WHO) offers an “analgesic ladder” guideline for stepwise medication intensification (9–12), pain management remains a challenge with heterogeneous opioid prescribing patterns among providers and approximately one-third of HNC patients still presenting to the emergency department with uncontrolled pain as a chief complaint. (13–16)

Challenges with pain control may be related to multifactorial clinical, cellular, and molecular mechanisms of pain, particularly in cancer patients. (17, 18) For example, endothelin-1, a vasoactive peptide with nociceptive effects, can be found in high levels in the tumor microenvironment of oral squamous cell carcinoma, and thereby may be a driver in the development of severe acute pain. (18) Inflammatory, neuropathic, and nociceptive pain can all be potentially stimulated during RT and present with certain attributes. For example, inflammatory pain is often described as a burning sensation while neuropathic pain is typically described as a numbness, tingling, shooting, or stabbing sensation. (19–22) The ability to differentiate or characterize acute pain features experienced by OC/OPC patients during active therapy may result in new information and added knowledge on how to effectively manage complex pain profiles.

While HNC pain-related studies exist in the literature, (17, 23–25) they are limited due to their broad scope (i.e., multi-symptom burden analysis) or focus on chronic pain and/or opioid dependency at a specified period of time after oncologic therapy. These studies also lack inclusion of pain descriptors and physiologic kinetics (i.e., the evolution of vital signs) which may elucidate underlying mechanisms of acute pain. To address this unmet need, the objective of this study was to provide an in-depth analysis on dynamic, acute pain profiles experienced by a modern, prospective OC/OPC cohort treated with curative RT. Temporal characterization of vital signs, provider-based treatment-related toxicity assessment (26), and analgesic prescription patterns during RT were also reported.

## Methods

### 2.1 Study Cohort

A prospective, electronic health record (EHR)-embedded registry has been developed at The University of Texas MD Anderson Cancer Center. All patients in this study had biopsy-proven squamous cell carcinoma (SCC) of the oral cavity (n= 120), oropharynx (n= 228), or metastatic SCC to the neck with an unknown primary (n=3), and were treated with curative (i.e., postoperative or definitive RT) from 2013 to 2021. RT prescriptions ranged from 60-70 Gy in 30-33 fractions, and RT modalities used included intensity-modulated radiotherapy (IMRT), volumetric modulated arc therapy (VMAT), or proton therapy. Patients could have multimodal therapy including upfront surgery and/or induction and concurrent chemotherapy. Patients were clinically staged using the AJCC (American Joint Committee on Cancer) 7^th^ edition (27). Exclusion criteria included patients of age < 18 years, those with a non-SCC OC/OPC diagnosis, and patients with metastatic disease. All patients must have completed at least 3 weekly see visit (WSV) assessments during their RT course and reported pain during one or more WSVs. This study was approved by the Institutional Review Board (IRB) at our institution.

### 2.2 Weekly Symptom and Vitals Assessment

EHR-based patient-reported symptom questionnaires at baseline (i.e., before RT) and during scheduled weekly see visits (WSVs) with treating clinicians in the department of Radiation Oncology were extracted. This questionnaire allows for incremental pain severity scoring on a 0 to 10 scale and includes qualitative parameters such as pain location, description, type, onset, and progression. Additional structured data on acute pain, prescribed analgesics, and WSV provider-based toxicity assessments were extracted from Brocade, our institutional, web-based electronic data capture system. (28) Extracted data for acute pain is HNC-site specific, therefore we focused on three main locations (i.e., mouth, throat, and skin) with associated pain descriptors (i.e., aching, sore, burning, sharp). WSV pain trajectories are denoted as baseline or WSV# (i.e., WSV6 signifies data from the 6^th^ WSV). Physician-graded mucositis and dermatitis are reported according to the National Cancer Institute Common Terminology Criteria for Adverse Events (version 5.0; CTCAE) (26). Overall treatment toxicity assessment is reported as being absent, mild, moderate, or severe.

Weekly physiologic parameters extracted from our EHR included weight (WT: Kg), blood pressure, and heart rate (HR, beats/min). The mean arterial pressure (MAP: mmHg) was calculated using the following formula: MAP = (systolic pressure + 2*diastolic pressure)/3. (29) Percent weight change was calculated using the following equation: 100*((current weight-baseline weight)/baseline weight).

### 2.3. Analgesic Assessment

Prescribed analgesics data and the cumulative analgesics used by patients were collected from our EHR and categorized into the following groups: 1) Topical analgesics (xyloxylin and lidocaine), 2) Non-opioids (anti-inflammatory drugs: NSAID, acetaminophen), 3) Weak opioids (codeine, hydrocodone, dihydrocodeine, and tramadol) (12, 30), 4) Strong opioids (morphine, hydromorphone, fentanyl, methadone, and oxycodone) (30) and 5) gabapentin. A similar approach to categorizing weak versus strong opioids based on WHO has been described by Gupta et al. (31) and colleagues (11, 32).

### 2.4 Statistical Analysis

Differences in weekly pain scores, weight, and vitals were assessed using one-way ANOVA tests. Tukey’s Honest Significant Difference (HSD) test was used to assess the significance of differences between pairs of group means. Trends in temporal pain scores and vital signs were assessed using a linear mixed-effect model (LMM) with random intercepts, accounting for associations between repeated measurements in the same subject. The study time period (i.e., during RT) was treated as a fixed effect and the subject as a random effect. Spearman’s rank correlation was used to calculate the correlation between changes in pain and vital signs. Differences in mucositis severity, dermatitis grades, treatment toxicity and opioid prescription were analyzed using the Chi-square test. AUC*pain* scores were calculated using previously described methods (33). Patients’ weekly pain scores were connected linearly and the area under the resulting curve is calculated and divided by the total area (maximum pain score × number of followed weeks). AUC pain metrics were then compared using Wilcoxon rank-sum test for binary variables and Kruskal-Wallis test for multilevel variables based on gender, age (>= 60 years old or <60 years old), primary tumor type, social habits (smoking, alcohol use and drug abuse), chemotherapy recipients, pre-RT surgery and pre-RT pain score.

All statistical tests were 2-sided and statistical significance was set at *P*< 0.05. Statistical analyses were performed using graph pad prism (version 8), JMP PRO 15 and R statistical software (version 4.0.3).

## Results

### 3.1 Cohort Characteristics

Patient, tumor, and treatment characteristics are outlined in Table 1. In a cohort of 351 OC/OPC patients [OC (n=120, 34%) and OPC (n=228, 66%)] treated with RT, the mean age at diagnosis was 58.5 years (range, 21 to 83 years, SD 10.8). The majority were White or Caucasian (n = 302, 86%) and male (n= 261, 74%). 32% and 31% of our cohort had a tonsil or base of tongue primary, respectively, and 59% had HPV-positive cancer. Most patients had a clinical stage with T1 or T2 (62%) disease and half had N2 (n=170, 49%) adenopathy. Surgery was used in 58% of the cohort, 14% received induction therapy, and 56% were treated with concurrent chemoradiation.

**Table 1:**
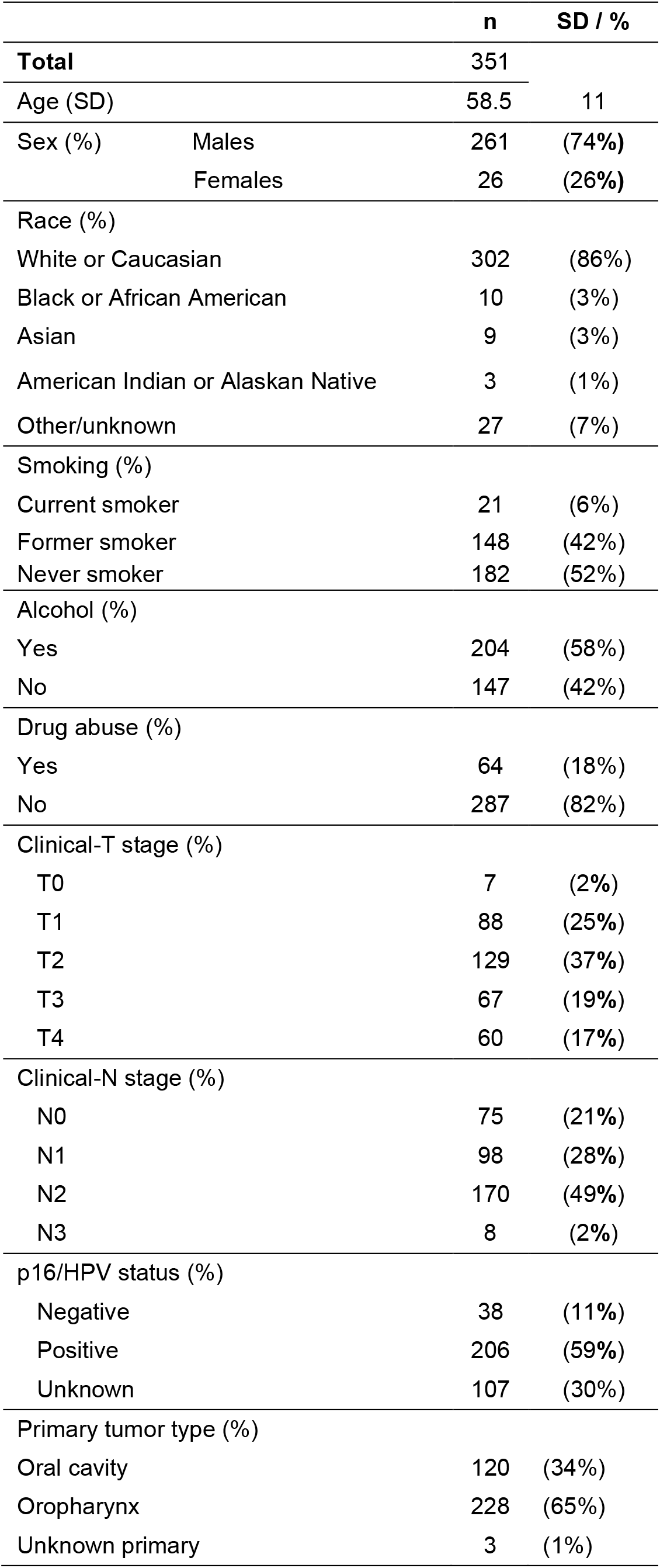
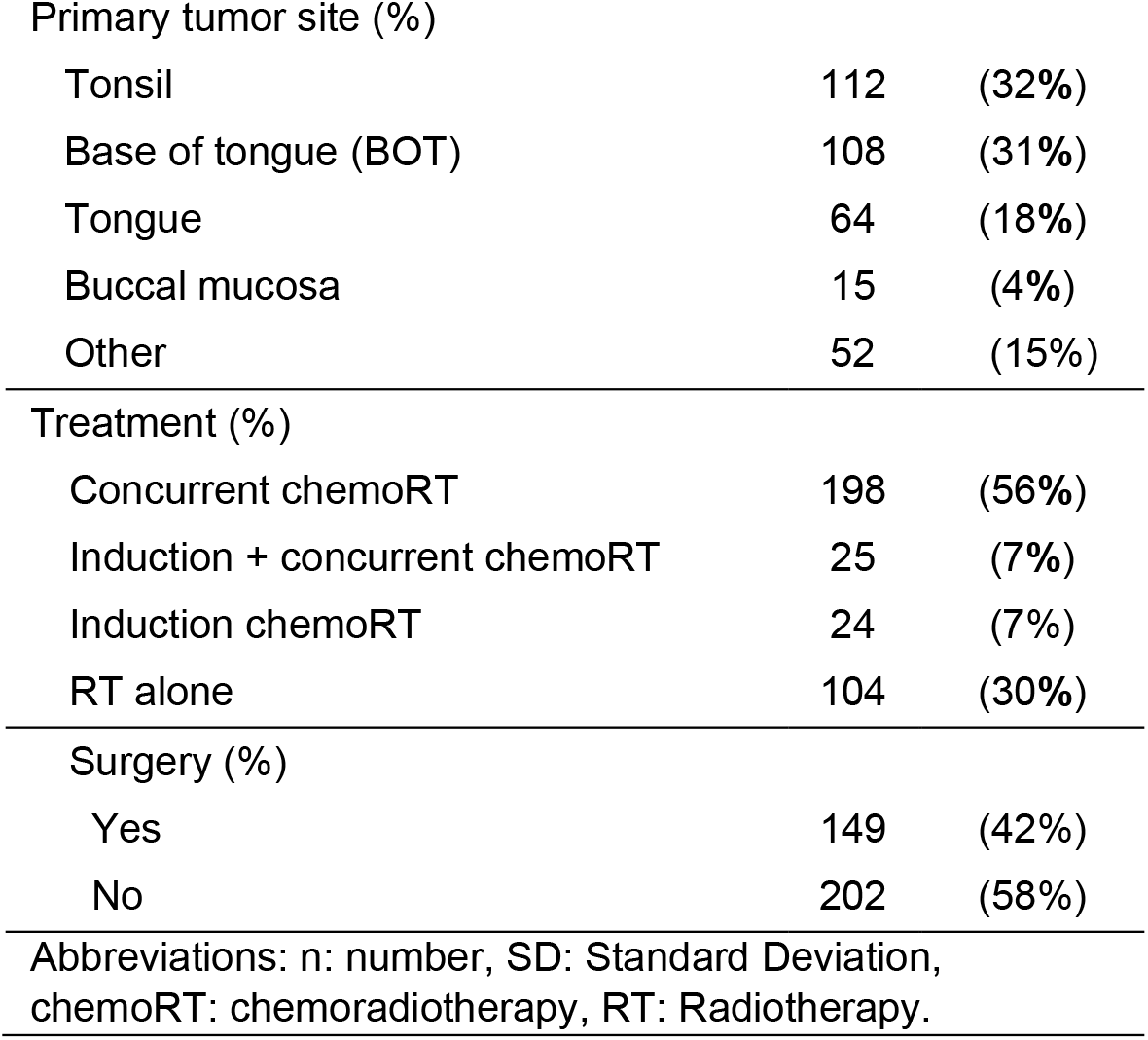
Patients characteristics

|  |  | n | SD / % |
| --- | --- | --- | --- |
| **Total** |  | 351 |  |
| Age (SD) |  | 58.5 | 11 |
| Sex (%) | Males | 261 | (74%) |
|  | Females | 26 | (26%) |
| Race (%) |  |  |  |
| White or Caucasian |  | 302 | (86%) |
| Black or African American |  | 10 | (3%) |
| Asian |  | 9 | (3%) |
| American Indian or Alaskan Native |  | 3 | (1%) |
| Other/unknown |  | 27 | (7%) |
| Smoking (%) |  |  |  |
| Current smoker |  | 21 | (6%) |
| Former smoker |  | 148 | (42%) |
| Never smoker |  | 182 | (52%) |
| Alcohol (%) |  |  |  |
| Yes |  | 204 | (58%) |
| No |  | 147 | (42%) |
| Drug abuse (%) |  |  |  |
| Yes |  | 64 | (18%) |
| No |  | 287 | (82%) |
| Clinical-T stage (%) |  |  |  |
| T0 |  | 7 | (2%) |
| T1 |  | 88 | (25%) |
| T2 |  | 129 | (37%) |
| T3 |  | 67 | (19%) |
| T4 |  | 60 | (17%) |
| Clinical-N stage (%) |  |  |  |
| N0 |  | 75 | (21%) |
| N1 |  | 98 | (28%) |
| N2 |  | 170 | (49%) |
| N3 |  | 8 | (2%) |
| p16/HPV status (%) |  |  |  |
| Negative |  | 38 | (11%) |
| Positive |  | 206 | (59%) |
| Unknown |  | 107 | (30%) |
| Primary tumor type (%) |  |  |  |
| Oral cavity |  | 120 | (34%) |
| Oropharynx |  | 228 | (65%) |
| Unknown primary |  | 3 | (1%) |

|  |  |  |
| --- | --- | --- |
| Primary tumor site (%) |  |  |
| Tonsil | 112 | (32%) |
| Base of tongue (BOT) | 108 | (31%) |
| Tongue | 64 | (18%) |
| Buccal mucosa | 15 | (4%) |
| Other | 52 | (15%) |
| Treatment (%) |  |  |
| Concurrent chemoRT | 198 | (56%) |
| Induction + concurrent chemoRT | 25 | (7%) |
| Induction chemoRT | 24 | (7%) |
| RT alone | 104 | (30%) |
| Surgery (%) |  |  |
| Yes | 149 | (42%) |
| No | 202 | (58%) |
| Abbreviations: n: number, SD: Standard Deviation, chemoRT: chemoradiotherapy, RT: Radiotherapy. |  |  |

### 3.2 Pain Assessment

Longitudinal pain profiles are reported in Table 2. A total of 341 (97.2%), 351 (99.7%), and 159 (45.3%) patients reported any pain score at baseline, WSV1, and WSV7. Mean (median) acute pain scores from baseline to WSV7 increased from 1.4 (0), 1.1 (0), 1.6 (1), 3 (2), 4(4), 4.2(4), 4.7(5), to 5.4(5), respectively. The overall change in pain intensity from WSV1 to WSV7 was significant (P<0.001) (Figure 1.a). Similarly, there was a significant difference in individual weekly pain scores (i.e., the change between one week and other weeks) except between WSV4 and WSV5. Throat and mouth pain were the most reported locations of pain, with 16% of patients reported throat/mouth pain with a median pain score of 3 (range, 0-10) (mean 3.4, SD=2.2) at baseline prior to starting RT. By WSV7, 63% and 78% of the cohort reported mouth and throat pain, respectively, with a median pain score of 6 (range, 1-10) (mean= 5.8, SD, 2.5). Reports of skin-related pain started at WSV2 (n=5, 1%) and showed a steady increase to 44% by WSV7. (Figure 1.b)

**Table 2:**
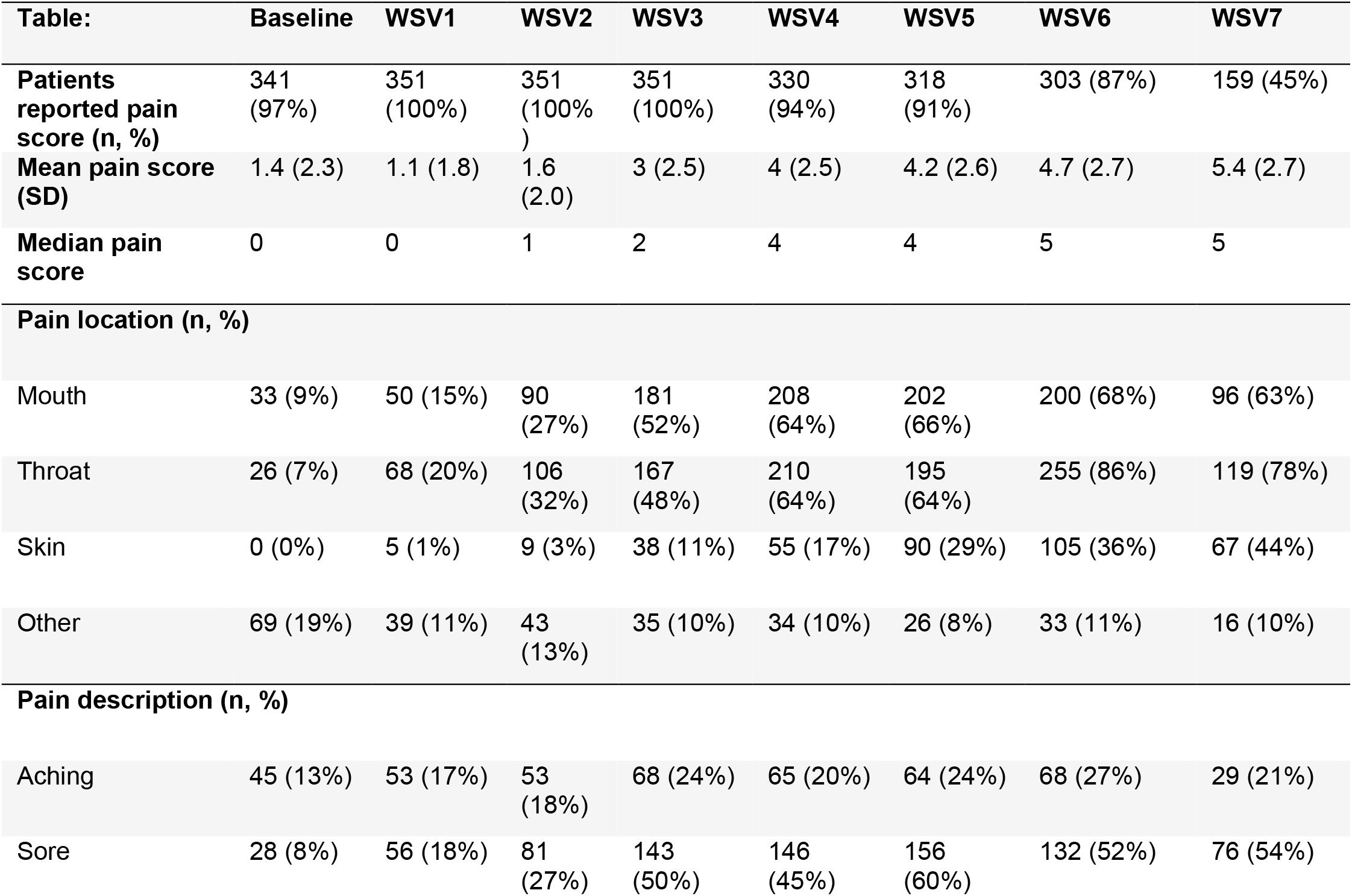
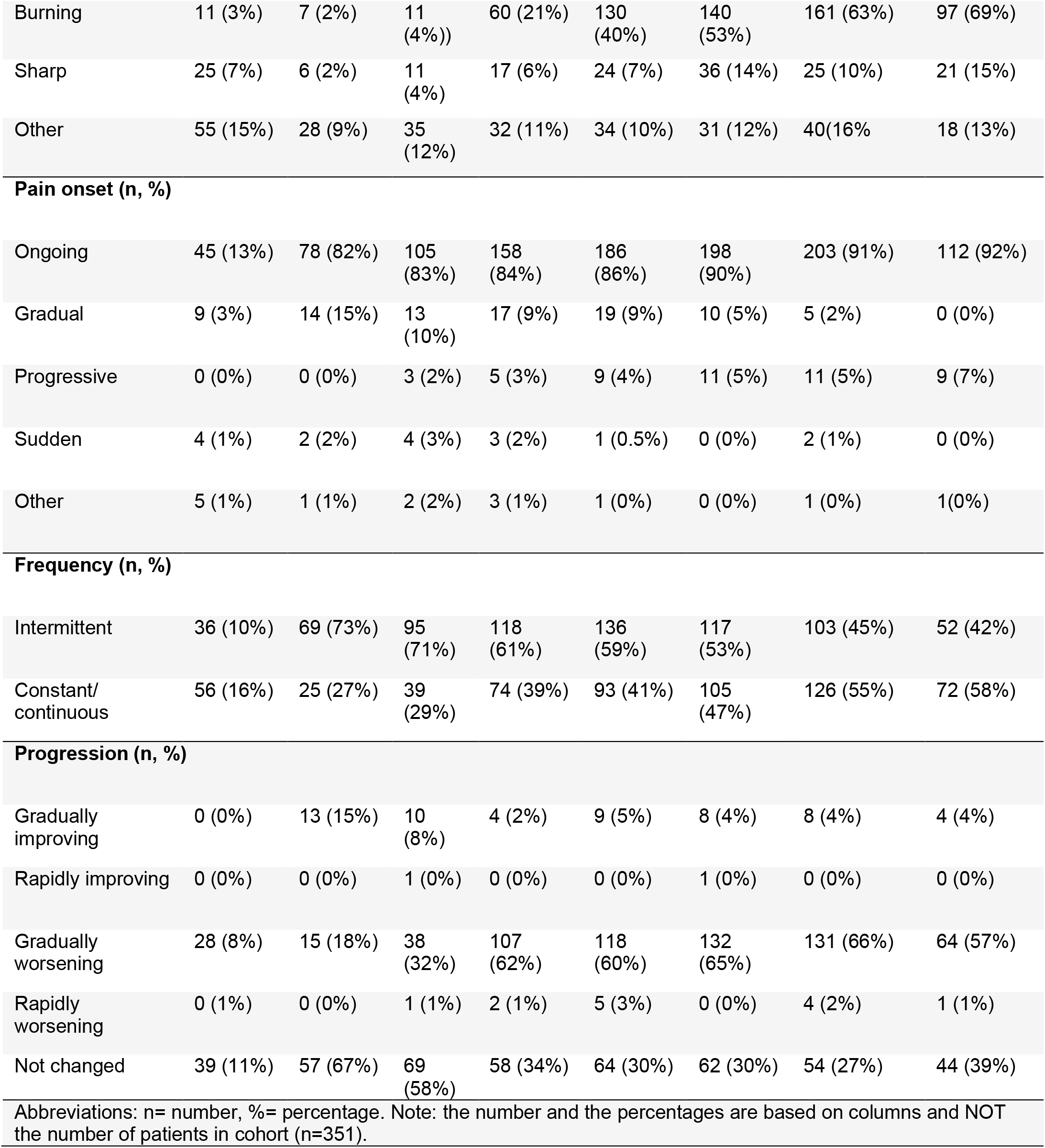
Patient reported pain trajectory and pain profiles during the weekly see visits (WSVs)

| Table: | Baseline | WSV1 | WSV2 | WSV3 | WSV4 | WSV5 | WSV6 | WSV7 |
| --- | --- | --- | --- | --- | --- | --- | --- | --- |
| **Patients reported pain score (n, %)** | 341 (97%) | 351 (100%) | 351 (100%) | 351 (100%) | 330 (94%) | 318 (91%) | 303 (87%) | 159 (45%) |
| **Mean pain score (SD)** | 1.4 (2.3) | 1.1 (1.8) | 1.6 (2.0) | 3 (2.5) | 4 (2.5) | 4.2 (2.6) | 4.7 (2.7) | 5.4 (2.7) |
| **Median pain score** | 0 | 0 | 1 | 2 | 4 | 4 | 5 | 5 |
| **Pain location (n, %)** |  |  |  |  |  |  |  |  |
| Mouth | 33 (9%) | 50 (15%) | 90 (27%) | 181 (52%) | 208 (64%) | 202 (66%) | 200 (68%) | 96 (63%) |
| Throat | 26 (7%) | 68 (20%) | 106 (32%) | 167 (48%) | 210 (64%) | 195 (64%) | 255 (86%) | 119 (78%) |
| Skin | 0 (0%) | 5 (1%) | 9 (3%) | 38 (11%) | 55 (17%) | 90 (29%) | 105 (36%) | 67 (44%) |
| Other | 69 (19%) | 39 (11%) | 43 (13%) | 35 (10%) | 34 (10%) | 26 (8%) | 33 (11%) | 16 (10%) |
| **Pain description (n, %)** |  |  |  |  |  |  |  |  |
| Aching | 45 (13%) | 53 (17%) | 53 (18%) | 68 (24%) | 65 (20%) | 64 (24%) | 68 (27%) | 29 (21%) |
| Sore | 28 (8%) | 56 (18%) | 81 (27%) | 143 (50%) | 146 (45%) | 156 (60%) | 132 (52%) | 76 (54%) |
| Burning | 11 (3%) | 7 (2%) | 11 (4%) | 60 (21%) | 130 (40%) | 140 (53%) | 161 (63%) | 97 (69%) |
| Sharp | 25 (7%) | 6 (2%) | 11 (4%) | 17 (6%) | 24 (7%) | 36 (14%) | 25 (10%) | 21 (15%) |
| Other | 55 (15%) | 28 (9%) | 35 (12%) | 32 (11%) | 34 (10%) | 31 (12%) | 40(16%) | 18 (13%) |
| Pain onset (n, %) |  |  |  |  |  |  |  |  |
| Ongoing | 45 (13%) | 78 (82%) | 105 (83%) | 158 (84%) | 186 (86%) | 198 (90%) | 203 (91%) | 112 (92%) |
| Gradual | 9 (3%) | 14 (15%) | 13 (10%) | 17 (9%) | 19 (9%) | 10 (5%) | 5 (2%) | 0 (0%) |
| Progressive | 0 (0%) | 0 (0%) | 3 (2%) | 5 (3%) | 9 (4%) | 11 (5%) | 11 (5%) | 9 (7%) |
| Sudden | 4 (1%) | 2 (2%) | 4 (3%) | 3 (2%) | 1 (0.5%) | 0 (0%) | 2 (1%) | 0 (0%) |
| Other | 5 (1%) | 1 (1%) | 2 (2%) | 3 (1%) | 1 (0%) | 0 (0%) | 1 (0%) | 1(0%) |
| Frequency (n, %) |  |  |  |  |  |  |  |  |
| Intermittent | 36 (10%) | 69 (73%) | 95 (71%) | 118 (61%) | 136 (59%) | 117 (53%) | 103 (45%) | 52 (42%) |
| Constant/<br>continuous | 56 (16%) | 25 (27%) | 39 (29%) | 74 (39%) | 93 (41%) | 105 (47%) | 126 (55%) | 72 (58%) |
| Progression (n, %) |  |  |  |  |  |  |  |  |
| Gradually improving | 0 (0%) | 13 (15%) | 10 (8%) | 4 (2%) | 9 (5%) | 8 (4%) | 8 (4%) | 4 (4%) |
| Rapidly improving | 0 (0%) | 0 (0%) | 1 (0%) | 0 (0%) | 0 (0%) | 1 (0%) | 0 (0%) | 0 (0%) |
| Gradually worsening | 28 (8%) | 15 (18%) | 38 (32%) | 107 (62%) | 118 (60%) | 132 (65%) | 131 (66%) | 64 (57%) |
| Rapidly worsening | 0 (1%) | 0 (0%) | 1 (1%) | 2 (1%) | 5 (3%) | 0 (0%) | 4 (2%) | 1 (1%) |
| Not changed | 39 (11%) | 57 (67%) | 69 (58%) | 58 (34%) | 64 (30%) | 62 (30%) | 54 (27%) | 44 (39%) |
| Abbreviations: n= number, %= percentage. Note: the number and the percentages are based on columns and NOT the number of patients in cohort (n=351). |  |  |  |  |  |  |  |  |

**Figure 1:**
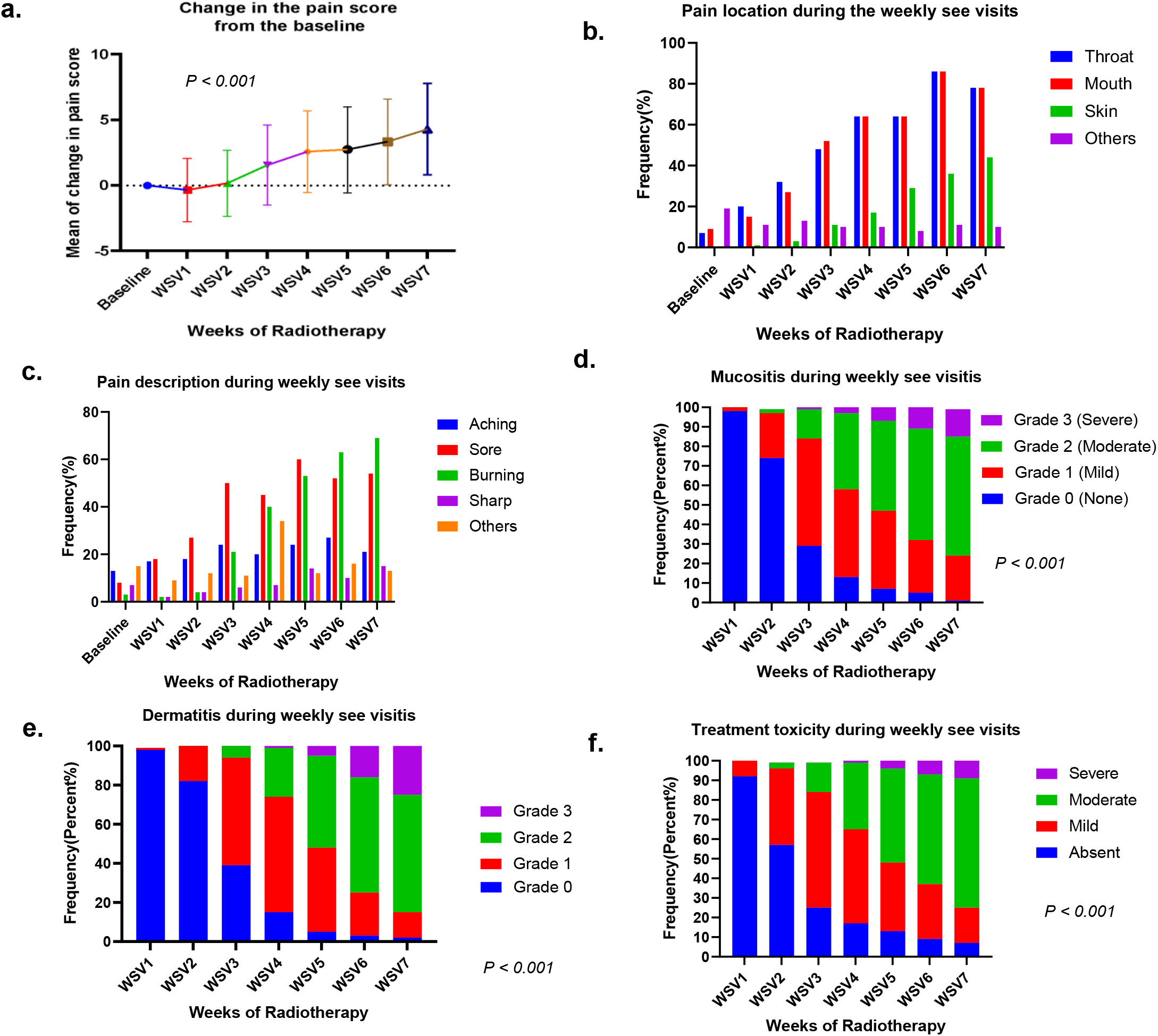
Acute pain profile and acute toxicities: **a**. Overall change of patient reported pain score (delta pain) showed significant increase in the mean pf pain score during the weeks of RT (WCVs) (*p<0.001*). **b**. Overall pain location reported in the weekly see visits. **c**. Overall pain description reported in the weekly see visits. **d**. Physician reported mucositis grades showed significant increase over the weeks of RT (*p<0.001*). **e**. Physician reported dermatitis grades showed significant increase over the weeks of RT (*p<0.001*). **f**. Physician reported overall treatment toxicity plan showed significant increase over the weeks of RT *(p<0.001*).

Multiple pain descriptors were experienced by patients over the course of RT (Figure 1.c). Aching pain remained relatively consistent through the WSVs (17%-27%), while soreness and burning sensations demonstrated the greatest temporal changes. For example, sore-specific pain, which was reported in 28 patients of the cohort and represented 8% of pain descriptors at baseline, peaked to 60% at WSV5 with a slight decrease to 54% by WSV7. Burning pain was less common during the baseline (3%) and the first 2 WSVs but began to increase significantly by WSV3 (21%) with a peak at end of treatment where 69% of patients reported this pain. Sharp pain showed a general positive trend over time with 2%, 7%, and 15% of the cohort experiencing this type of pain during WSV1, WSV4, and WSV7, respectively.

Additional longitudinal data on pain onset, frequency, and progression were analyzed. Most patients expressed their pain as ongoing between weeks (82-92%) with a higher propensity for gradual onset over sudden onset. Pain was often felt to be intermittent (WSV1, 73%) early on during RT with a gradual shift towards being more constant during the latter part of RT (WSV7, 58%). This coincided with a gradual worsening of pain over time (WSV1, 18% to WSV7, 57%) with minimal gradual/rapid improvement in pain despite delivered analgesic measures.

In efforts to condense temporal information related to pain trajectories into a single metric, AUC*pain* scores were calculated for the entire cohort (Median 16.0% (IQR (9.3–23.0)) and for specific subgroups. Significant difference in AUCpain scores in subgroups based on gender (median male 16.5% (IQR 9.5-24.5), female 13.8% (IQR 8.5-19.9), *p=0.048*), primary cancer type (median oral cavity 14.5% (IQR 7.5-20.0), oropharynx 17.5% (10.4-25.0), *p=0.002*), primary tumor site (BOT 19.8% (IQR 12.5-25.6), Tonsils 14.0% (IQR 8.5-21.1), others 14% (IQR 9.0-21.6), *p= 0.003*), surgery (none 17.5% (IQR 11.0-25.0), post 12.5% (6.0-20.0), *p<0.001*), drug abuse (No 14.5 % (IQR 9-22), Yes 20.8 (IQR 12.4-26.3), p=0.003 and pre-RT pain score (non-mild (score 0-4) 20.3% (12.1-30.0), moderate-severe (score 5-10) 15.5% (9.0-22.3), *p=0.012*) while no significant difference were detected in subgroups based on chemotherapy received (*p=0.468*), age (*p=0.385*), smoking (*p=0.234*) or alcohol history (*p=0.315*). (Figure 2)

**Figure 2:**
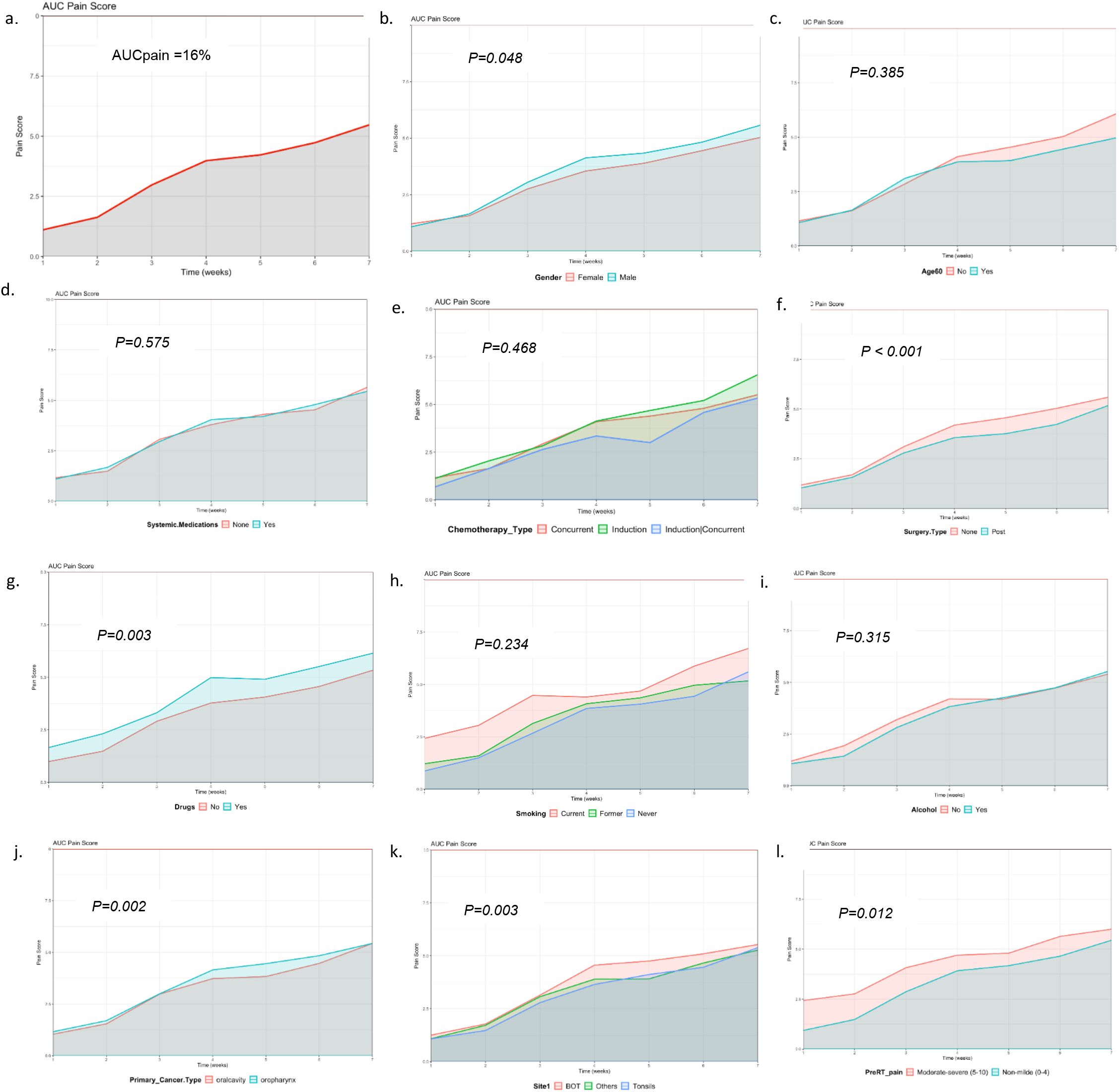
AUCpain scores. **a**. Overall pain AUC curve (16%). **b**. AUCpain scores based on gender, male group had significant higher AUC pain curve than female (*p=0.048*). **c**. AUCpain scores based on age, no significant difference between < 60 and >60 years old (*p=0.385*). **d**. AUCpain scores of systemic medication (chemotherapy) use showed no significant difference (*p=0.575*). **e**. AUCpain scores based on chemotherapy type showed no significant difference (p=0.468). **f**. AUCpain scores based on pre-RT surgery, patients had RT post-surgery (Post) had significant lower pain curve than patients did not have surgery (None) (*p<0.001*). **g**. AUCpain scores based on drug abuse, patients had history of drug abuse had significant higher pain curve than patients did not (No) (*p=0.003*). **h**. AUCpain scores based on smoking status showed no significant difference (p=0.234). **i**. AUCpain scores based on alcohol status showed no significant difference (*p=0.315*). **j**. AUCpain scores based on primary cancer type, oropharynx group had significant higher AUC pain curve than oral cavity (*p=0.002*). **k**. AUCpain scores based on primary cancer site, Base of tongue group had significant higher AUC pain curve than tonsils and others (p=0.003). l. AUCpain scores based on pre-RT pain scores, group had non-mild pain score (0-4) had lower AUC curve than group

### 3.3 Provider-based Objective Toxicity Assessments

Weekly mucositis and dermatitis showed significant increase in severity over time (*P*<0.001). There was a sharp decline in grade 0 mucositis from WSV2 (74%) to WSV3 (29%) with progressive severity peaking at WSV7 where rates of grade 0, 1, 2, and 3 mucositis were 1%, 23%, 61%, 14%, respectively (Figure 1.d). Similar trajectories in dermatitis grading were noted with grade 2+ dermatitis increasing from 26%, 75%, and 85% in WSVs 4, 6, and 7 (P<0.001; Figure 1.e). Provider-based overall treatment toxicity grading is based on careful assessment of a variety of factors including physiologic changes, pain control, dermatitis, and mucositis changes. From a clinician’s perspective, most patients exhibited moderate toxicity to treatment by WSV7 (66%) while severe toxicity was only reported in 9% of the cohort (Figure 1.f).

### 3.4 Physiologic Changes During RT

A significant decline of 7.1% in percent body weight for the entire cohort was seen by the end of RT (95%CI, 9.96-8.17; *P<0.0001*) (Figure 3.a). This correlated with an increasing rate of feeding tube insertions during RT, with 4% of patients having a feeding tube at WSV1 compared to 13% and 29% during weeks 5 and 7 (*P*<0.001; Figure 3.b). A mean arterial pressure (MAP) drop of 6.8 mmHg and corresponding increase in mean HR by 10.7 beats/min (95%CI, 7.56-13.82; *P<0.0001*) was seen from WSV1 to 7 (*P*<0.001, Figure 3.c-d).

**Figure 3:**
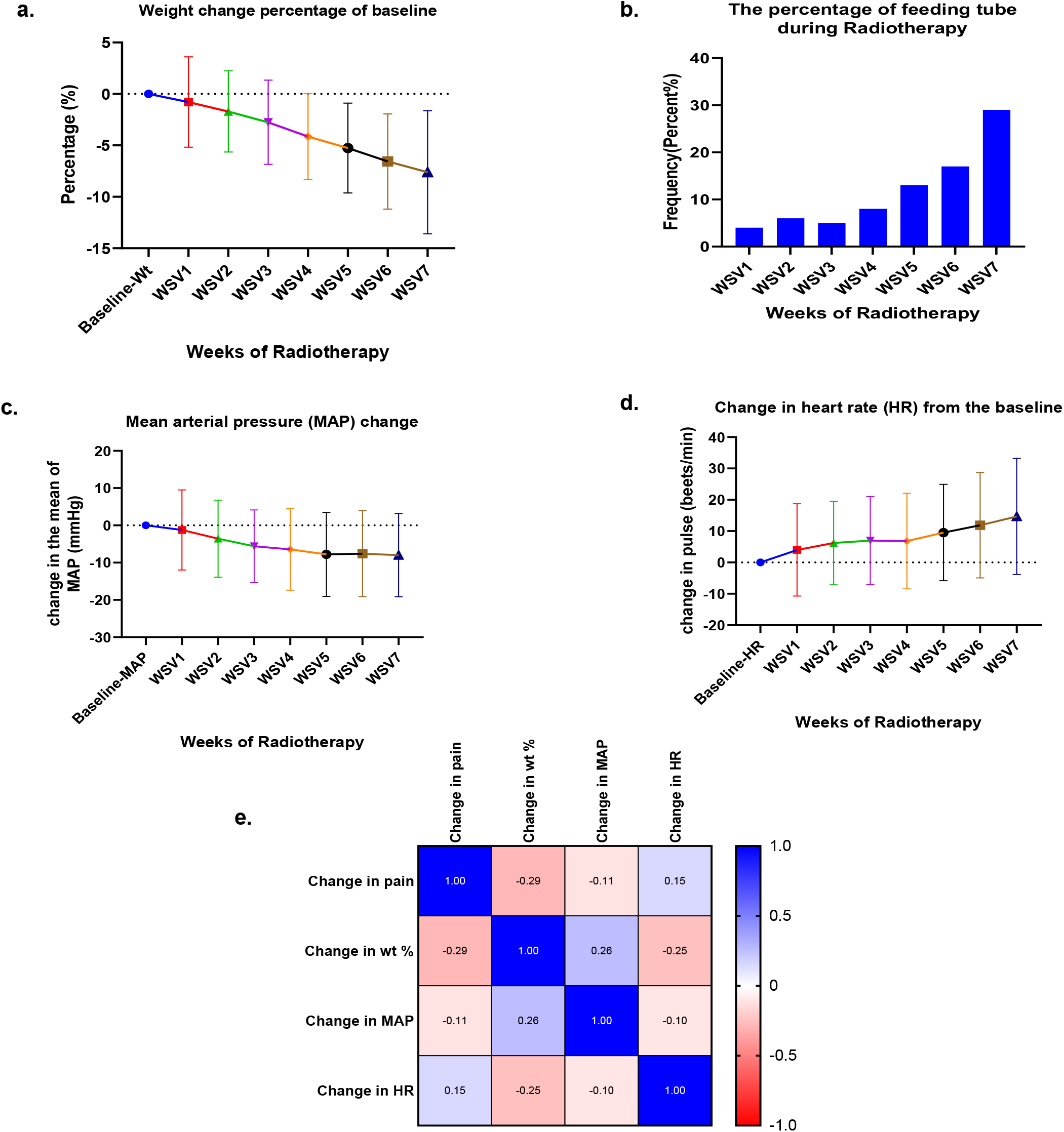
Physiologic changes: **a**. Temporal overall change in the mean of body weight change during the weeks of RT. **b**. Frequency of feeding tube used through the weekly see visits of RT. **c**. Temporal change in the mean of mean arterial pressure (MAP) change during the weekly see weekly see visits. **d**. Temporal change in heart rate (HR) change during the weekly see visits. **e**. Spearman r correlation matrix showing correlation between the change in pain and the change in vital signs.

Using a Spearman r correlation matrix (Figure 2e), a positive association with time was noted for pain score changes (95%CI, 0.7–0.8; *P*< 0.001) and HR change (95%CI, 1.20 –1.82); *P<0.001*) while a negative correlation was seen with weight change (95%CI, −1.29 to −1.10; *P<0.001*) and MAP change (95%CI, −6.29 to −4.90; *P*<0.001). Additionally, a positive correlation between pain and HR (r= 0.15, 95%CI, 0.11-0.19; *P<0.001*) as well as between weight and MAP (r= 0.26, 95%CI, 0.22-0.30; *P<0.001*) were observed. A negative correlation was detected between pain and weight (r= −0.29, 95%CI, −0.32 to −0.25; *P<0.001*), pain and MAP (r= −0.12, 95%CI, −0.15 to −0.07; *P<0.001*), weight and HR (r= −0.25, 95%CI, −0.29 to −0.21; *P<0.001*) and MAP and HR (r= −0.10, 95%CI, −0.14 to −0.06; *P<0.001*) (Figure 3.e).

### 3.5 Analgesics/Opioid Prescription Patterns

Analgesics data analysis revealed a temporal increase in the cumulative frequency of different analgesics used by patients through the weeks of RT (Figure 4.a). Topical analgesics and weak opioids are the most commonly used analgesics throughout RT with increase in the frequency of topical anesthetics from 22% in WSV1 reaching the peak on WSV5 (70%) with slight decrease in WSV6 (64%), similarly increase in the frequency of weak opioids use from 21% in WSV1 up to 69% and 68% in WSV5 and WSV6 respectively, while strong opioids use starts with 3% in WSV1 till reaching the peak on WSV6 (34%) with decrease to 22% in WSV7. Furthermore, non-opioids analgesics and Gabapentin use starts with 11 % and 13% respectively in WSV1 and reaches the peak by WSV5 with 15% and 27% respectively and then decrease to 4% and 13% in WSV7. The frequency of strong opioids’ prescription showed a statistically significant increase through the WSVs, with an average 45% increase from WSV1 (10%) to the end of RT (average of week 6 and 7) (55%), while gradual decrease in weak opioids’ prescription through the weeks of RT from 90% in WSV1 to 53% in WSV7 (P=<0.0001). (Figure 4.b).

**Figure 4:**
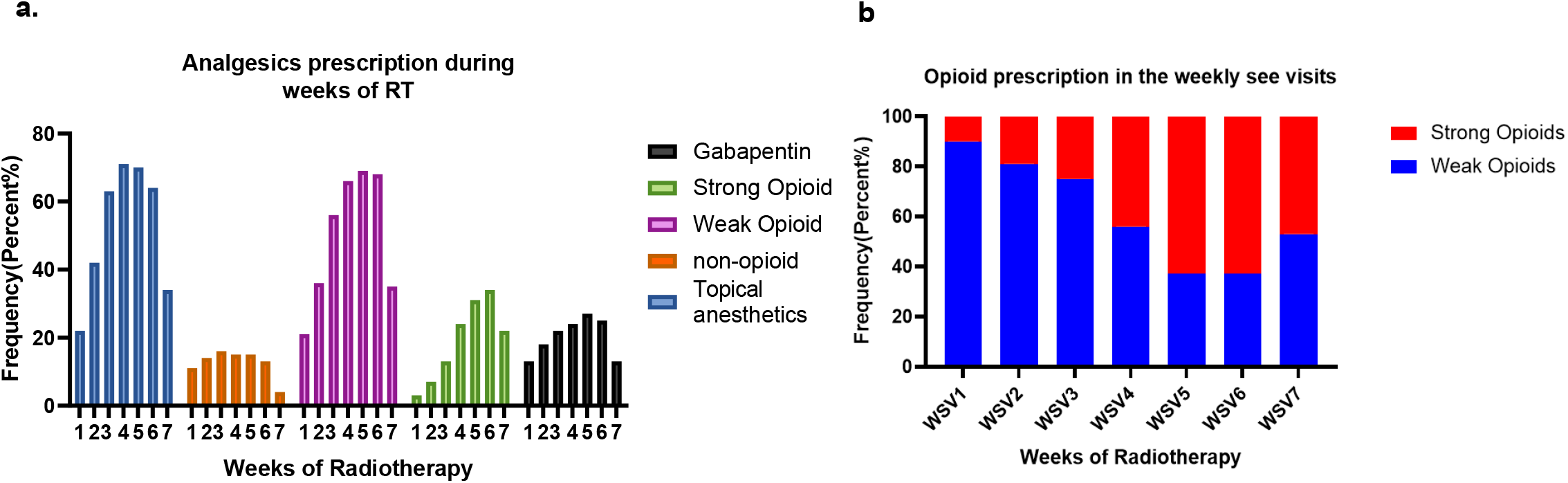
**a**. Temporal cumulative frequency of analgesics used by patients during the weekly see visits (WSVs) of radiotherapy. **b**. The frequency of weak opioids versus strong opioids prescribed during the WSVs shows significant difference in opioids prescription (*p<0.001*).

## Discussion

Pain is a significant baseline and acute toxicity experienced by OC/OPC patients who are treated with RT. The etiology of pain can be related to the primary disease itself or other progressive acute radiation-induced toxicities such as oral mucositis and dermatitis. (34–38) Despite continued advancements in RT delivery methods and systemic therapy options, recent randomized HNC clinical trials such as RTOG 1016 still show high rates of acute toxicities like grade 3-4 oral mucositis affecting 42-46% of the cohort. (39) In this same trial, acute pain was reported as a limited “all terms” rating of grade 3-4 pain (15%), which represents only a part of the cohort’s experience and lacks any temporal information or insight on how acute pain was managed. Therefore, the objective of our study was to perform a comprehensive analysis on the temporal characterization of longitudinal sequelae of RT including acute pain and relevant physiologic changes and provider-based toxicity assessments using a modern, prospective EHR-embedded HNC registry. We found significant increase in the cumulative patient reported acute pain scores (*p<0.001*) and physician reported acute oral mucositis and dermatitis grades through the course of RT, being mouth, throat, and skin burning pain as the most commonly pain sites and description reported. A significant correlation between pain severity and poor nutritional and physiological status of patients represented with significant weight loss, MAP drop, and increase in HR from the baseline to the end of RT was found.

We previously reported increase in pain severity as 1 out of 7 different patient-reported symptom trajectories throughout the course of RT (6-7 weeks) (p= 0.051) using a validated patient reported outcome measures called MD Anderson Symptom Inventory–Head and Neck Module (MDASI-HN) on a prospective longitudinal HNC cohort study (25). Our current study builds upon those previous results by exploring AUCpain metrics and granular AP profiles to preserve temporal pain data, analyzing objective acute toxicity assessment, and studying the physiologic burden of AP on patients’ health and outcome. Additionally, we previously developed a new quantitative measure of overall symptom burden AUC*symptom* throughout the period of RT (33), and in this study we used similar approaches to describe pain trajectories in this cohort as overall symptom and in subgroups based on age, sex, social habits, treatment recipient and primary tumor type. Significant differences were found in subgroups stratified according to gender, primary tumor site, surgery, and drug use history. Although the impact of chemotherapy on symptom burden has been identified in previous literature (4, 33, 40), this effect was not detected in our cohort. Male patients and patients who had history of drug abuse, pre-RT surgery, and oropharynx primary tumor type in base of the tongue had higher AUC metrics, while no significant difference was detected in subgroups stratified based on age, smoking, alcohol use and chemotherapy. These metrics may be useful for future toxicity prediction model building.

Different characteristics of acute pain (19, 20, 41–44), acute mucositis (36, 37, 45), and acute dermatitis (46) have been described in the literature, all of which were stimulated by RT, mainly due to stimulation of inflammatory cells (47) and release of different cytokines in response to radiation (48, 49) inducing inflammatory reaction and stimulation of nociceptors (21) resulting in pain (45), our data supported these results and, we have shown an increase in the frequency of burning pain, mucositis and dermatitis throughout the course of treatment.

Epstein et al demonstrated that HNC patients complain of head and neck pain during RT which impacts quality of life despite ongoing pain management.(41) Our results support Epstein et.al. and exhibited ongoing increase in pain intensity in OC/OPC during RT despite an increase in analgesics use. Mucositis and dermatitis are dose dependent, showed increase in the severity with the gradual increase in radiation dose and fraction (45). Our study showed a gradual increase in the severity of mucositis and dermatitis overtime with an increase in radiation done and fraction.

To holistically analyze pain management of HNC patients undergoing RT, full data of analgesics prescribed, the time of these prescriptions during RT (which week), and accurate reporting and collection of patient medication use is important. This will provide useful information as to how pain is managed as patients undergo RT at their weekly visits. Our study demonstrated increase in analgesics-prescription, particularly strong opioids during weeks of RT, while significant pain at the end of treatment with minimal/no improvement in pain severity was reported reflecting a continual unmet need for pain control optimization during RT. A previous study showed that following the WHO analgesics ladder resulted in patients expressing better quality of life and undisturbed activities. It also showed that most patients undergoing cancer treatment quickly escalated along the prescription ladder to requiring strong opioids for adequate pain management (9, 11). Schaller et al. performed a descriptive study of pain management in HNC patients during RT revealed that strong opioids represented the majority of analgesics used for moderate to severe pain management, with alleviation of pain in patients receiving structured use of opioids, and severe pain was association with patients presenting with high grades of mucositis (2+) (32).

Although we performed a longitudinal prospective study using a comprehensive data analysis of a large cohort of OC/OPC patients, limitations of the study include missing some pain and medication use data, since some patients didn’t describe pain or precise use of prescribed medications in detail. Analysis of other confounding factors affecting pain experience such as tumor staging, medical history (e.g., chronic diseases) is needed. Furthermore, studying the effect of pain on the quality of life was not investigated, which will give better indication of the general impact of AP and acute toxicities on patient’s outcome. Further prospective data collection to improve recording of patient medication use data is also needed to improve accuracy of correlation of analgesic use with pain control. This study relied on prescriptions provided as this data was not available, however, not all patients may have been taking prescriptions exactly as prescribed.

Our future goal is to use the results of this study as preliminary data for the development of statistically robust, AI toxicity prediction models and pain-based algorithmic treatment paradigms which will be discussed, separately.

In conclusion, our results demonstrated a significant temporal increase in the severity of AP and treatment-related acute toxicities over time throughout the course of RT in OC/OPC patients and the ongoing need for pain control in this population. The severity of AP change is associated with poor nutritional and health status during receiving RT. AUCpain metrics and the significant impact of different clinical and demographic features on AP burden could be used in the future to develop a predictive algorithm for pain management optimization.

## Data Availability

All data produced in the present study are available upon reasonable request to the authors.

## Notes

### Competing Interest Statement

The authors have declared no competing interest.

### Funding Statement

This study was funded by Principal Investigator, 8 months, Pain-directed Pragmatic Automation of Informatics-based Novel Frameworks for Reducing Radiotherapy-associated Effects (PAINFREE), 5K12CA088084, Paul Calabresi K12 Scholars Program, 9/1/2020-8/31/2022

### Author Declarations

Ethics committee/IRB [2020-1231] of University of Texas, MD Anderson Cancer Center gave ethical approval for this work.

